# Diffusion MRI-based measures of neurite microstructure associate with future risk of Alzheimer’s Disease

**DOI:** 10.1101/2025.04.07.25325401

**Authors:** Sasha Hakhu, Andrew Hooyman, Jennapher Lingo VanGilder, Sydney Y. Schaefer, Scott C. Beeman, Alzheimer’s Disease Neuroimaging Initiative

## Abstract

**INTRODUCTION:** Early detection of Alzheimer’s disease (AD) is crucial for intervention, but traditional MRI and cognitive assessments may miss pre-symptomatic changes. Advanced diffusion MRI (dMRI) methods, such as Neurite Orientation Dispersion and Density Imaging (NODDI), show promise in identifying early brain changes.

**METHODS:** We analyzed 65 cognitively unimpaired older adults (25 APOE-e4 carriers, 40 non-carriers) from the ADNI3 dataset. NODDI’s neurite density index (NDI) and orientation dispersion index (ODI), volumetric MRI and cognitive performance (MoCA) were analyzed in key brain regions like the hippocampus, fusiform gyrus, and entorhinal cortex. Statistical analyses included linear regression and t-tests, with FDR correction.

**RESULTS:** NDI differed significantly between carriers and non-carriers and correlated with MoCA scores. ODI differed only in the CA1 hippocampal subfield. Volumetric MRI measures showed no group differences.

**DISCUSSION:** NODDI metrics, particularly NDI, could help detect early APOE-e4-related microstructural changes, while traditional volumetric MRI measures remain uninformative at early stages.

## 1. Introduction

Early disease identification and intervention are critical for treating Alzheimer’s disease (AD). Therapies targeting amyloid and tau slow disease progression, potentially extending the patient’s quality of life^1^; however, these treatments are most effective earlier on^2^. Advances in imaging techniques have significantly enhanced our ability to detect changes leading up to AD. Positron emission tomography (PET) uses radiotracers to measure AD-linked abnormalities like glucose metabolism^3,4^, amyloid plaque deposition and tau neurofibrillary tangles ^5–7^. Amyloid-PET visualizes plaques, which disrupt neuronal communication and trigger inflammation, leading to neurodegeneration before cognitive decline^5^. Tau-PET tracks tau tangles that disrupt neuronal transport systems, resulting in cell death^6^. Other tools such as neuropsychological assessments, blood biomarkers, and structural magnetic resonance imaging (MRI) can monitor symptoms and pathology^5,8–10;^ but only reflect macroscopic changes occurring *after* cognitive deficits. They fail to capture subtle, early microstructural alterations in brain tissue preceding neurodegeneration and symptom onset. A non-invasive method is needed to directly measure neurodegeneration by quantifying actual neuronal degradation, characteristic of neurodegenerative diseases like AD.

Diffusion magnetic resonance imaging (dMRI) offers an avenue for early and accurate AD screening ^11^ by detecting microstructural changes^12–14^ that precede symptomatic AD. Although dMRI itself is not a novel technique, recent advancements in acquisition protocols and biophysical modelling have dramatically expanded its utility in quantification of neuropathological processes^15^, particularly those involving neurodegeneration^15,16^. dMRI non-invasively measures the diffusion-driven displacement of water in spaces like intra/extra-cellular spaces and neurites to infer tissue microstructural features quantitively (e.g., cell size or neurite density)^17^; thus capturing diffusion patterns reflecting underlying structures. For example, water in white matter moves freely axially but is restricted radially, producing anisotropy. In gray matter, diffusion is more uniform thus creating isotropic patterns. Multi-shell dMRI captures varied tissue microenvironments, and biophysical models quantify diffusion corresponding to microstructure (e.g., neurite density/orientation). It provides a non-invasive, repeatable method for distinguishing between pathological and healthy tissue microstructure, with applications in detecting AD ^18,19^. Specifically, neurite orientation dispersion and density imaging (NODDI)^20^ models the diffusion signal by compartmentalizing volume fractions into "free water," "intra-neurite," and "extra-neurite" spaces. NODDI quantifies neurite structural integrity, where higher neurite density index (NDI) values indicate greater neurite density, and lower orientation dispersion index (ODI) values reflect greater coherence.

The APOE-e4 allele increases AD risk^21,22^ for early- and late-onset cases. ^23,24^ Studies show strong correlations between structural degradation and APOE-e4 ^7,25,26^ suggesting neurite density loss may be an early marker of disease pathology. ^7,25–29^ PET imaging detects amyloid plaques^5^ and tau proteins^5,30^, important biomarkers of AD. However, due to its high costs, limited availability, reliance on radioactive tracers, and lack of direct reporting on actual neurodegeneration (loss of cell bodies and/or neurites), other non-invasive methods for early detection and monitoring of neurodegeneration are needed. dMRI is non-invasive, accessible, cost-effective and provides a direct measure of neurodegeneration based on microstructural measures, making it complementary to plasma biomarkers for early detection, especially in pre-symptomatic individuals.

We tested whether dMRI-measured neurite structural integrity was lower in cognitively unimpaired APOE-e4 carriers than non-carriers. Neuroimaging data from the Alzheimer’s Disease Neuroimaging Initiative (ADNI) database (adni.loni.usc.edu) were analysed using NODDI-derived NDI and ODI calculated in prospectively-identified regions of interest, including the fusiform gyrus, entorhinal cortex, and hippocampal subfields (cornu ammonis CA1, CA2-3)^31–36^. NODDI-based metrics were correlated with the Montreal Cognitive Assessment (MoCA). Standard volumetric MRI measures were evaluated to determine whether diffusion metrics offered unique insights. We hypothesized that APOE-e4 carriers would have lower NDI (i.e., reduced neurite density) and higher ODI (i.e., increased neurite dispersion) values in the fusiform gyrus, entorhinal cortex, and hippocampal subfields than non-carriers.

Given the greater significance of left hemispheric hippocampal changes in AD development^22,35^, we hypothesized lower NDI and higher ODI values in the left hemisphere of carriers. Lastly, we hypothesized that MoCA scores would significantly correlate with dMRI-based metrics, but not with standard T1-based volume measures, given that microstructural changes like alterations in neurite density and orientation dispersion, likely occur much earlier in the disease process before substantial volume loss is apparent^12–14^.

## 2. Methods

### 2.1. Participants

Sixty-five cognitively unimpaired older adults were analyzed in this study to obtain brain dMRI and neuropsychological testing data. Mean age overall was 69.5± 5.5 (range = 62 to 90, 45F). The dataset used in preparation of this study was obtained from the Alzheimer’s Disease Neuroimaging Initiative (ADNI3, 2017) ^37^ database (adni.loni.usc.edu), consisting of cognitively unimpaired APOE-e4 carriers (n=25) and non-carriers (n=40). ‘Carriers’ were defined as having a least one e4 allele. The carrier and non-carrier groups were not significantly different in terms of age (p = 0.22).

### 2.2. Neuroimaging data acquisition

Data used in the preparation of this article were obtained from the Alzheimer’s Disease Neuroimaging Initiative (ADNI) database (adni.loni.usc.edu). The ADNI was launched in 2003 as a public-private partnership, led by Principal Investigator Michael W. Weiner, MD. The primary goal of ADNI has been to test whether serial magnetic resonance imaging (MRI), positron emission tomography (PET), other biological markers, and clinical and neuropsychological assessment can be combined to measure the progression of mild cognitive impairment (MCI) and early Alzheimer’s disease (AD).

Data used in this study were acquired using the ADNI-3 advanced protocol ^37^ on a 3T scanner. The following data were processed in this study: (i) MPRAGE T1-w image (TR = 2300 ms, TI = 900 ms) and (ii) multi-shell diffusion data with three shells: b = 500,1000, 2000 s/mm^2^ (TR/TE 3300/71 ms, 112 total diffusion weighted directions) and thirteen interleaved b = 0 s/mm^2^ values.

### 2.3. Neuroimaging data processing

Data were pre-processed using FSL ^38^ and MRtrix ^39^. The raw diffusion data underwent - noise correction, susceptibility distortion correction (Synb0 ^40^), motion and eddy current-induced distortion correction ^41^ and bias field correction.

The NODDI model was then applied to the diffusion data (MATLAB toolbox ^42^) to obtain neurite density index (NDI) and orientation dispersion index (ODI), which are both scaled from 0 to 1. NDI is a measure of neurite volume fraction (a value of 1 suggesting a denser population of neurites), while ODI is a measure of neurite coherence (a value of 1 suggesting greater dispersion of neurites). Functionally speaking, NDI reflects the density of neurite packing while ODI reflects the degree of aligned or scattered neurites in space.

Following this, brain-extracted T1 and b0 images underwent linear image co-registration^43^, followed by a non-linear transformation ^44^ to the Montreal Neurological Institute (MNI) standard brain atlas ^45^. For this study specifically, regions-of-interest (ROIs) were acquired from the Harvard-Oxford cortical and subcortical structural atlases ^46^ and non-linearly transformed to an older brain atlas ^47^ (to address issues such as age-related ventricular displacement, etc.). ROI-based mean values and standard deviations for NDI and ODI were extracted and analyzed. These regions of interest (fusiform gyrus, entorhinal cortex and hippocampal subfields) were selected a priori based on previous demonstration of their crucial role in memory and other cognitive functions^31–36,48–51^.

Additionally, standard volumetric calculations were calculated by using these segmented brain ROIs, overlaying them onto T1-weighted images, extracting the volume measures for each region (using FSL), and then comparing the volumetric differences between carriers and non-carriers.

### 2.4. Cognitive assessment

Scores from the Montreal Cognitive Assessment (MoCA) ^37^ were also analysed for the participants included in this study. Although the participants were classified as cognitively unimpaired within the ADNI dataset based on extensive neuropsychological evaluation, examining MoCA scores allowed for the investigation of potential associations between global cognitive scores, APOE-e4 carrier status, and microstructural changes at the neurite level.

### 2.5. Statistical analysis

Statistical analyses were performed using R version 4.2.2 ^52^. Linear regression models were employed to compare NDI, ODI, and volumetric measurements in the left (L) and right (R) hemispheric regions of the brain between APOE-e4 carrier and non-carrier groups. Brain regions analysed included the fusiform gyrus, entorhinal cortex, and hippocampal subfields (cornu ammonis CA1, CA2-3, CA4). The focus on hippocampal subfields is justified due to their established association with AD ^19,53–55^. By analysing the left and right regions individually rather than looking at them as a summed total, we aimed to capture lateralized effects that may be relevant to the pathology ^21,35,56,57^. ROIs were spatially normalized through warping and co-registration (in the preprocessing steps) to align template ROI masks to each participant’s coordinate space and address individual differences in brain size and shape.

A linear regression model was used to examine the effect of APOE-e4 status on various brain region metrics. APOE-e4 status (carrier vs. non-carrier) was modelled as the independent variable, while the mean NDI and volume values of different ROIs and cognitive scores (MoCA) were treated as dependent variables. To assess the significance of the differences between the carrier and non-carrier groups, t-tests were performed between MRI metrics for each region and cognitive score between groups. A false discovery rate (FDR) correction step was applied to account for multiple comparisons. Box plots and scatter plots were generated to visualize the differences between the groups.

Additionally, because MoCA scores did not vary based on carrier status (see Results), groups were collapsed across carrier status to evaluate the extent to which MoCA scores varied as a function of NDI and ODI values and standard volumetric measures in specific regions of interest. We examined the sensitivity of these diffusion metrics relative to volumetric methods based on cognitive measures and APOE-e4 status. Volumetric and NDI measures were standardized to z-scores for comparison, and linear regression models were used to analyse how measurements of brain regions, such as NDI and volume, affect MoCA scores, while controlling for age and sex.

## 3. Results

Figures 1, 2 and 3 show NDI, ODI, and standard volumetric calculations respectively, for the left and right regions of interest. Region-specific results and participant demographics are summarized in Table 1. Results demonstrated a significant effect of APOE-e4 carrier status (carrier vs. non-carrier) on NDI values in the left fusiform gyrus (β = 0.015, 95% CI = [0.002; 0.02], p = 0.02), right fusiform gyrus (β = 0.018, 95% CI = [0.003; 0.03], p = 0.02) (Fig. 1a), left entorhinal cortex (β = 0.018, 95% CI = [0.002; 0.03], p = 0.04) and right entorhinal cortex (β = 0.018, 95% CI = [0.001; 0.03], p = 0.03) (Fig. 1b). Furthermore, NDI values were significantly different within the left hippocampal subfield regions CA1 (β = 0.03, 95% CI = [0.005; 0.06], p = 0.02) (Fig. 1c) and CA2-3 (β = 0.03, 95% CI = [0.005; 0.05], p = 0.02) (Fig. 1d) between carriers and non-carriers, although the right hippocampus subfield regions showed no significant differences in NDI values based on carrier status (right CA1: β = 0.01, 95% CI = [-0.01; 0.03], p = 0.27; right CA2-3: β = -0.002, 95% CI = [-0.04; 0.04], p = 0.9).

**Figure 1.**
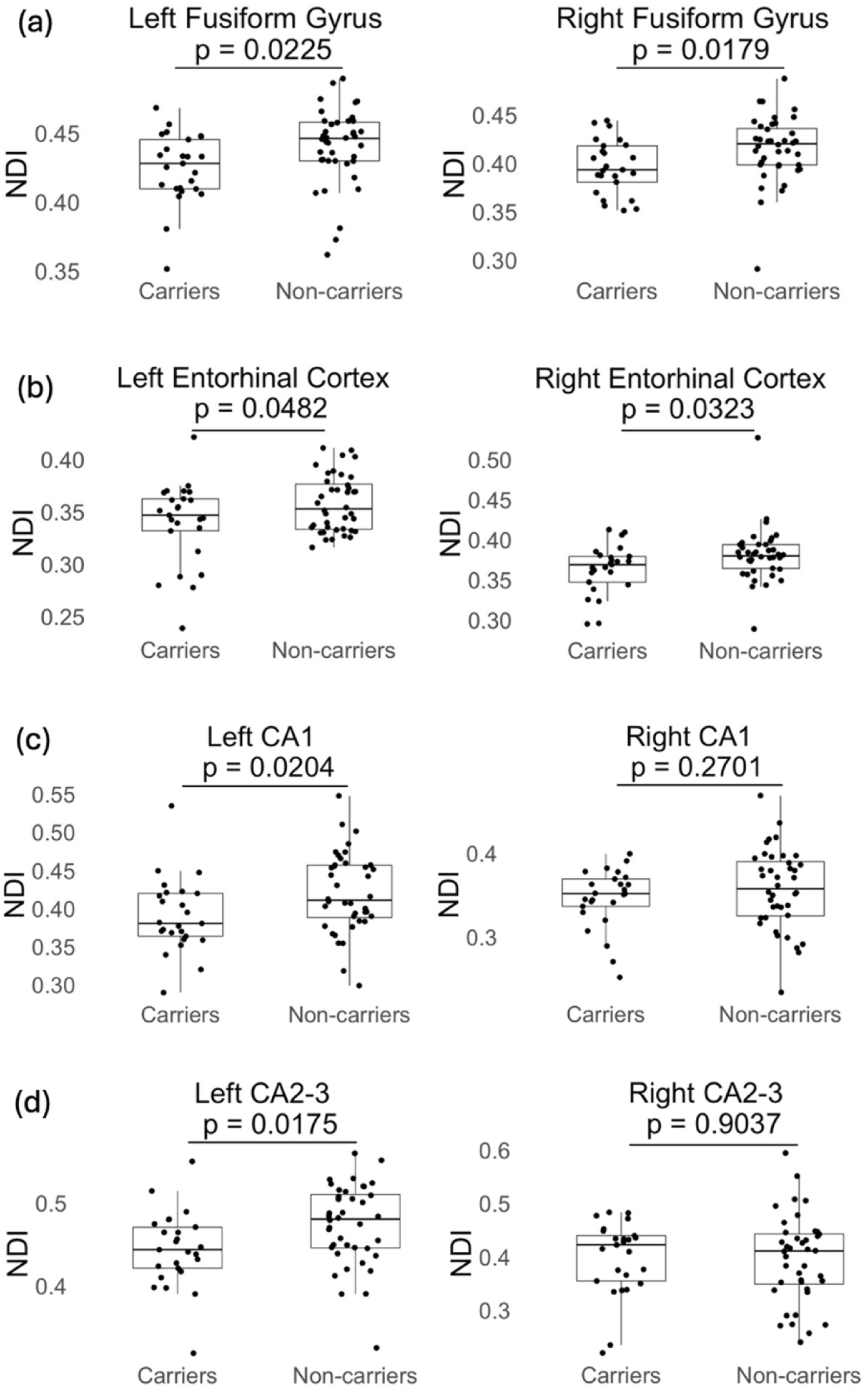
Neurite density index (NDI) values in the left and right (a) fusiform gyrus, (b) entorhinal cortex, (c) CA1 and (d) CA2-3 regions amongst APOE-e4 carriers and non-carriers.

**Figure 2.**
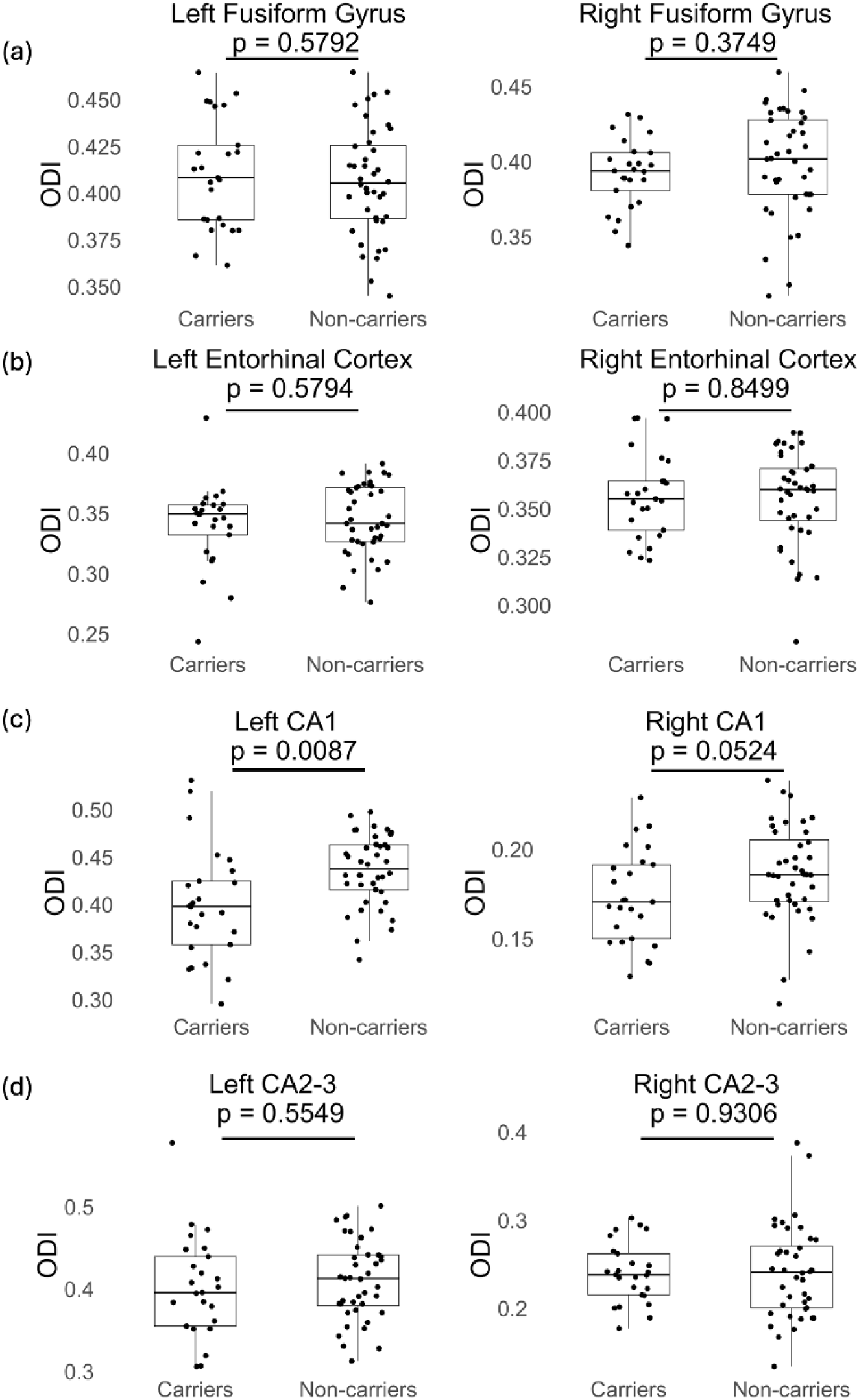
Orientation dispersion index (ODI) values in the left and right (a) fusiform gyrus, (b) entorhinal cortex, (c) CA1 and (d) CA2-3 regions amongst APOE-e4 carriers and non-carriers.

**Figure 3.**
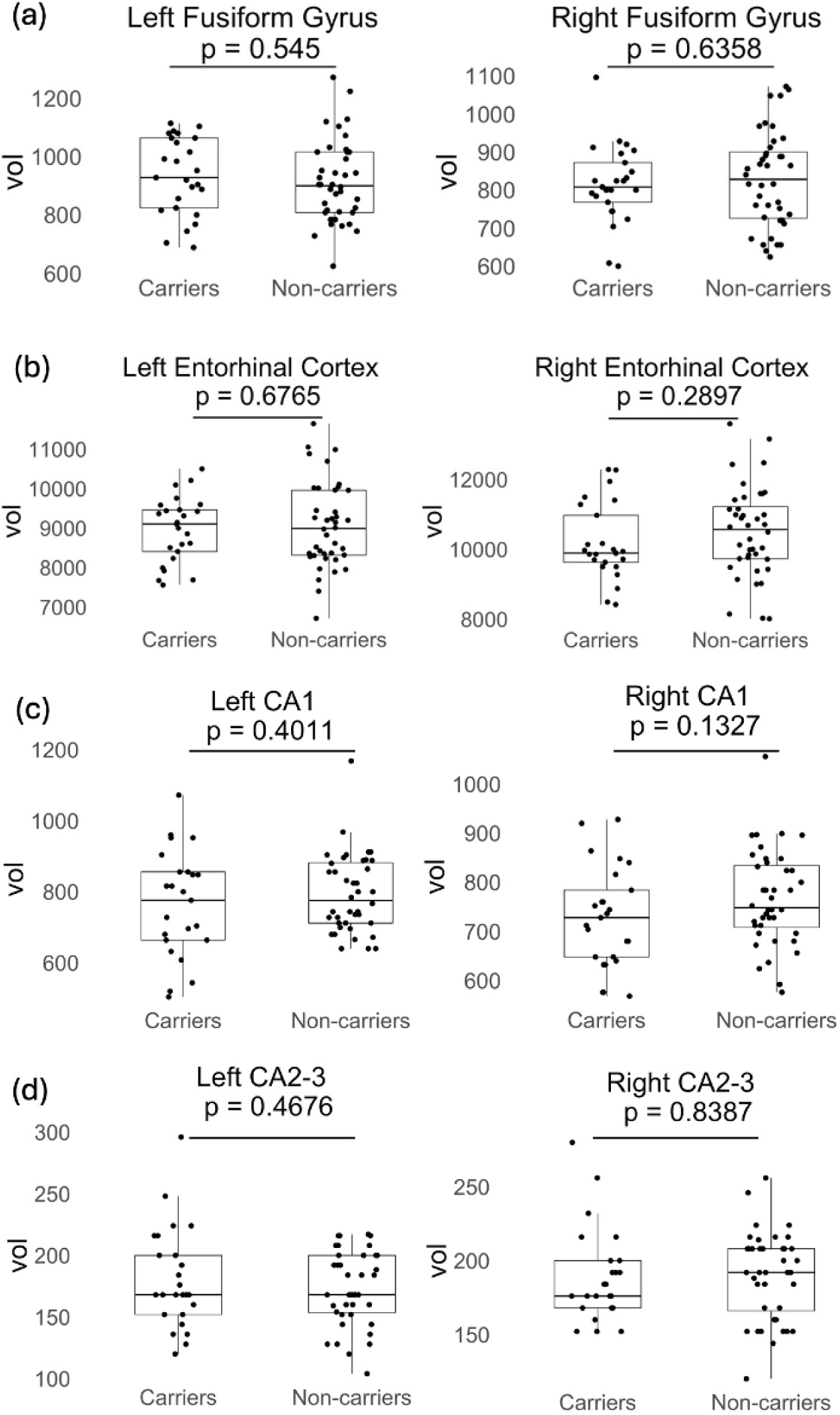
Volumetric analysis values (vol) in the left and right (a) fusiform gyrus, (b) entorhinal cortex, (c) CA1 and (d) CA2-3 regions amongst APOE-e4 carriers and non-carriers.

**Table 1.** Neuroimaging results of left (L) and right (R) ROIs for cognitively unimpaired older adults stratified by carrier status: APOE-e4 carriers (e4+) and non-carriers (e4-).

|  | ROI | e4- (n=40) | e4+ (n=25) | p-value<br>(FDR) |
| --- | --- | --- | --- | --- |
| **NDI** | **L fusiform gyrus** | 0.44 ± 0.03 | 0.42 ± 0.03 | 0.022* |
|  | **R fusiform gyrus** | 0.42 ± 0.03 | 0.39 ± 0.03 | 0.018* |
|  | **L entorhinal cortex** | 0.36 ± 0.03 | 0.33 ± 0.03 | 0.048* |
|  | **R entorhinal cortex** | 0.38 ± 0.03 | 0.36 ± 0.03 | 0.032* |
|  | **L CA1** | 0.42 ± 0.03 | 0.39 ± 0.05 | 0.020* |
|  | **R CA1** | 0.36 ± 0.05 | 0.35 ± 0.04 | 0.270 |
|  | **L CA2-3** | 0.47 ± 0.05 | 0.44 ± 0.05 | 0.017* |
|  | **R CA2-3** | 0.40 ± 0.08 | 0.40 ± 0.07 | 0.903 |
| **ODI** | **L fusiform gyrus** | 0.41 ± 0.03 | 0.41 ± 0.03 | 0.578 |
|  | **R fusiform gyrus** | 0.40 ± 0.04 | 0.40 ± 0.02 | 0.420 |
|  | **L entorhinal cortex** | 0.34 ± 0.03 | 0.34 ± 0.03 | 0.563 |
|  | **R entorhinal cortex** | 0.36 ± 0.02 | 0.36 ± 0.02 | 0.853 |
|  | **L CA1** | 0.44 ± 0.04 | 0.40 ± 0.06 | 0.008* |
|  | **R CA1** | 0.19 ± 0.03 | 0.17 ± 0.03 | 0.052* |
|  | **L CA2-3** | 0.41 ± 0.05 | 0.40 ± 0.06 | 0.532 |
|  | **R CA2-3** | 0.24 ± 0.05 | 0.24 ± 0.03 | 0.937 |
| **Volume (vol)**<br>**(in mm<sup>3</sup>)** | **L fusiform gyrus** | 912 ± 140 | 932 ± 130 | 0.545 |
|  | **R fusiform gyrus** | 828 ± 125 | 814 ± 103 | 0.636 |
|  | **L entorhinal cortex** | 9062 ± 1008 | 8962 ± 900 | 0.678 |
|  | **R entorhinal cortex** | 10498 ± 1200 | 10182 ± 1100 | 0.299 |
|  | **L CA1** | 793 ± 110 | 764 ± 150 | 0.401 |
|  | **R CA1** | 766 ± 97 | 727 ± 102 | 0.133 |
|  | **L CA2-3** | 174 ± 30 | 180 ± 40 | 0.468 |
|  | **R CA2-3** | 190 ± 29 | 189 ± 31 | 0.839 |

\* indicates statistically significant result ( $p < .05$ ); L: left; R: right

ODI values for these regions displayed significant differences between carriers and non-carriers only in the left CA1 (β = 0.037, 95% CI = [0.013; 0.06], p = 0.008) and right CA1 (β = 0.013, 95% CI = [-0.0001; 0.03], p = 0.052) (Fig. 2c) subfields. All other ROIs were not significantly different between groups: left fusiform gyrus (β = -0.004, 95% CI = [-0.019; 0.010], p = 0.578), right fusiform gyrus (β = 0.006, 95% CI = [-0.009; 0.02], p = 0.420) (Fig. 2a), left entorhinal cortex (β = 0.005, 95% CI = [-0.011; 0.021], p = 0.563); right entorhinal cortex (β = - 0.001, 95% CI = [-0.013; 0.011], p = 0.853) (Fig. 2b); left CA2-3 (β = 0.009, 95% CI = [-0.018; 0.036], p = 0.532) and, right CA2-3 (β = 0.0009, 95% CI = [-0.022; 0.025], p = 0.937) (Fig. 2d).

In contrast, no difference in carrier status was observed when analyzed using the standard volumetric measurement for the left fusiform gyrus (β = -21.03, 95% CI = [-90.99; 48.94], p = 0.54), right fusiform gyrus (β = 13.65, 95% CI = [-46.29; 73.58], p = 0.64) (Fig. 3a), left entorhinal cortex (β = 99.93, 95% CI = [-407.85; 607.70], p = 0.68) or right entorhinal cortex (β = 315.85, 95% CI = [-300.31; 932.01], p = 0.29) (Fig. 3b), left CA1 (β = 29.01, 95% CI = [- 34.94; 92.96], p = 0.40), right CA1 (β = 39.17, 95% CI = [-11.39; 89.74], p = 0.13) (Fig. 3c), left CA2-3 (β = -6.92, 95% CI = [-24.51; 10.67], p = 0.47), right CA2-3 (β = 1.60, 95% CI = [-13.83; 17.03], p = 0.84) (Fig. 3d).

Figure 4 show that MoCA scores were not significantly different between carriers and non-carriers (β = 0.51, 95% CI = [-0.88; 1.90], p = 0.51), which was expected given that all participants were asymptomatic. Thus, the groups were collapsed across carrier status to evaluate the extent to which MoCA scores varied as a function of dMRI and standard volumetric measures in the specific regions of interest (Figs. 5 and 6). NDI correlated with MoCA scores in left entorhinal cortex (raw p= 0.05), right entorhinal cortex (raw p=0.001), left fusiform gyrus (raw p= 0.02) and right CA2-3 (raw p= 0.02) regions. Neither ODI nor standard volumetric measures showed a strong association with MoCA scores. The increased sensitivity of NDI relative to standard volumetric methods to global cognition as well as APOE-e4 carrier status is illustrated in Table 2.

**Figure 4.**
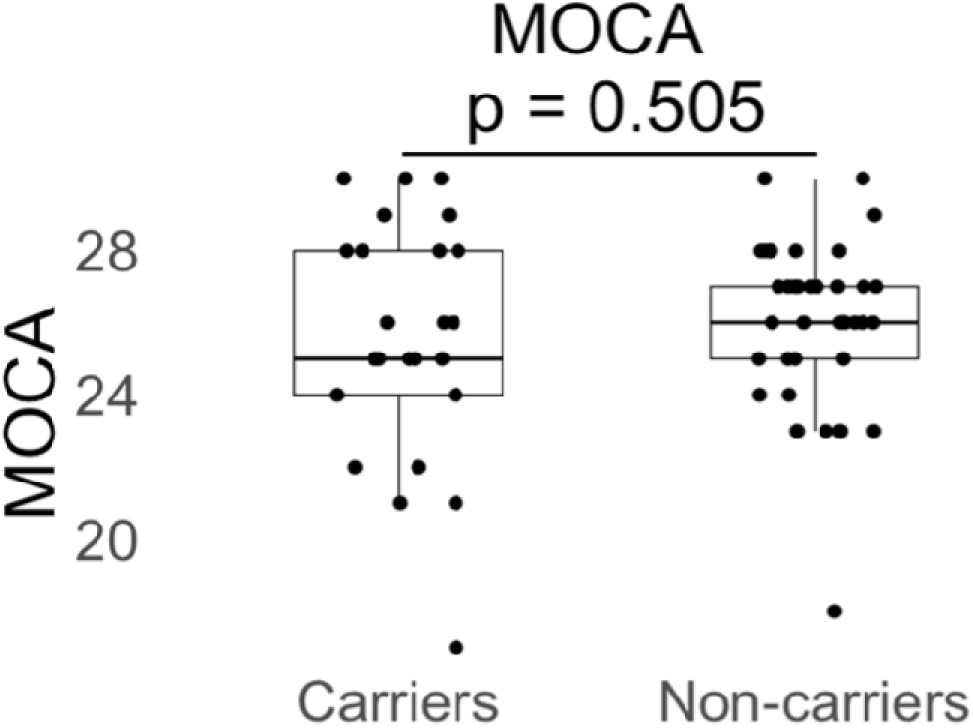
Montreal Cognitive Assessment (MoCA) test scores amongst APOE-e4 carriers and non-carriers.

**Figure 5.**
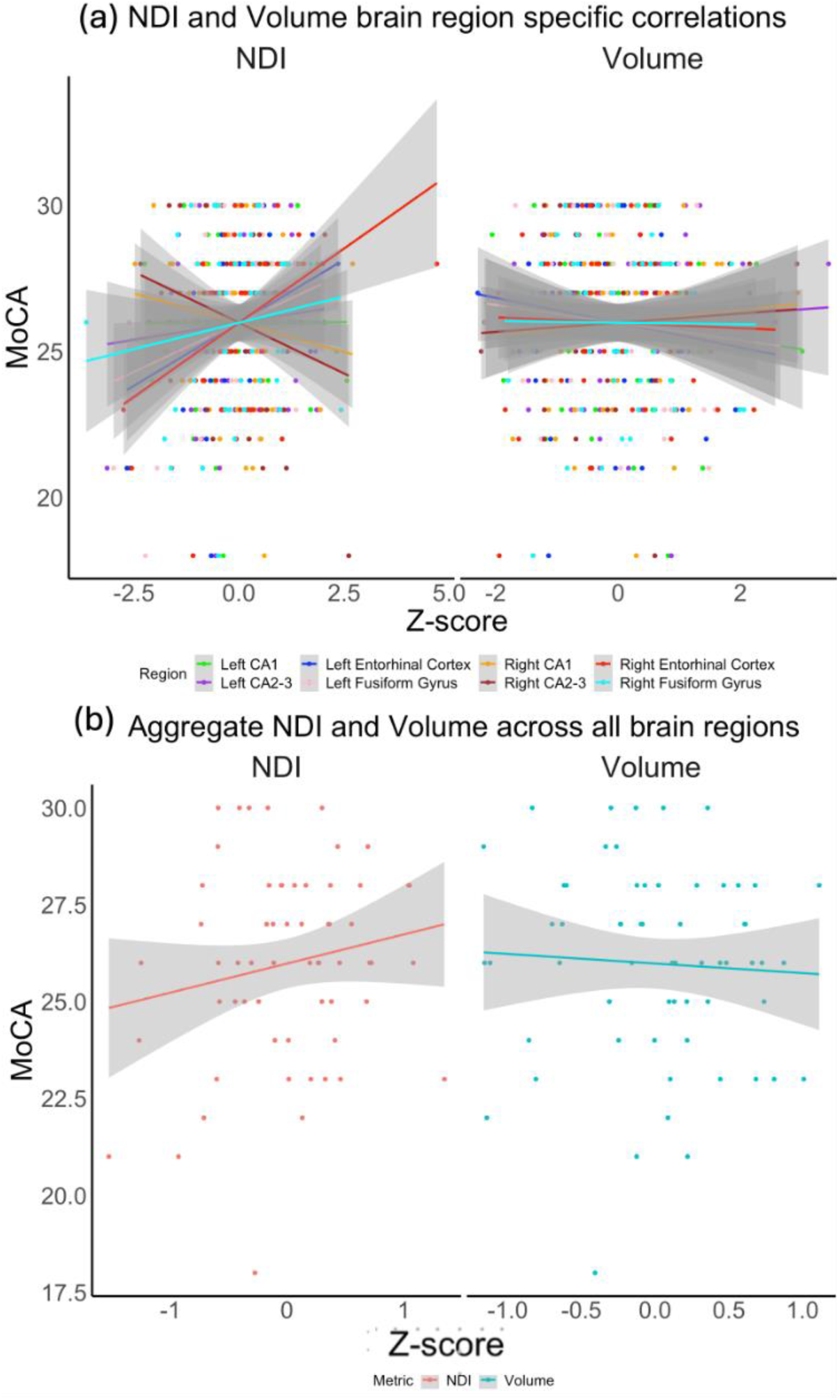
(a) Plots showing correlation between NDI and MoCA scores in the left and right CA1, CA2-3, entorhinal cortex and fusiform gyrus. (b) Aggregate NDI and volume measures for each participant across all brain regions regressed to MoCA score.

**Figure 6.**
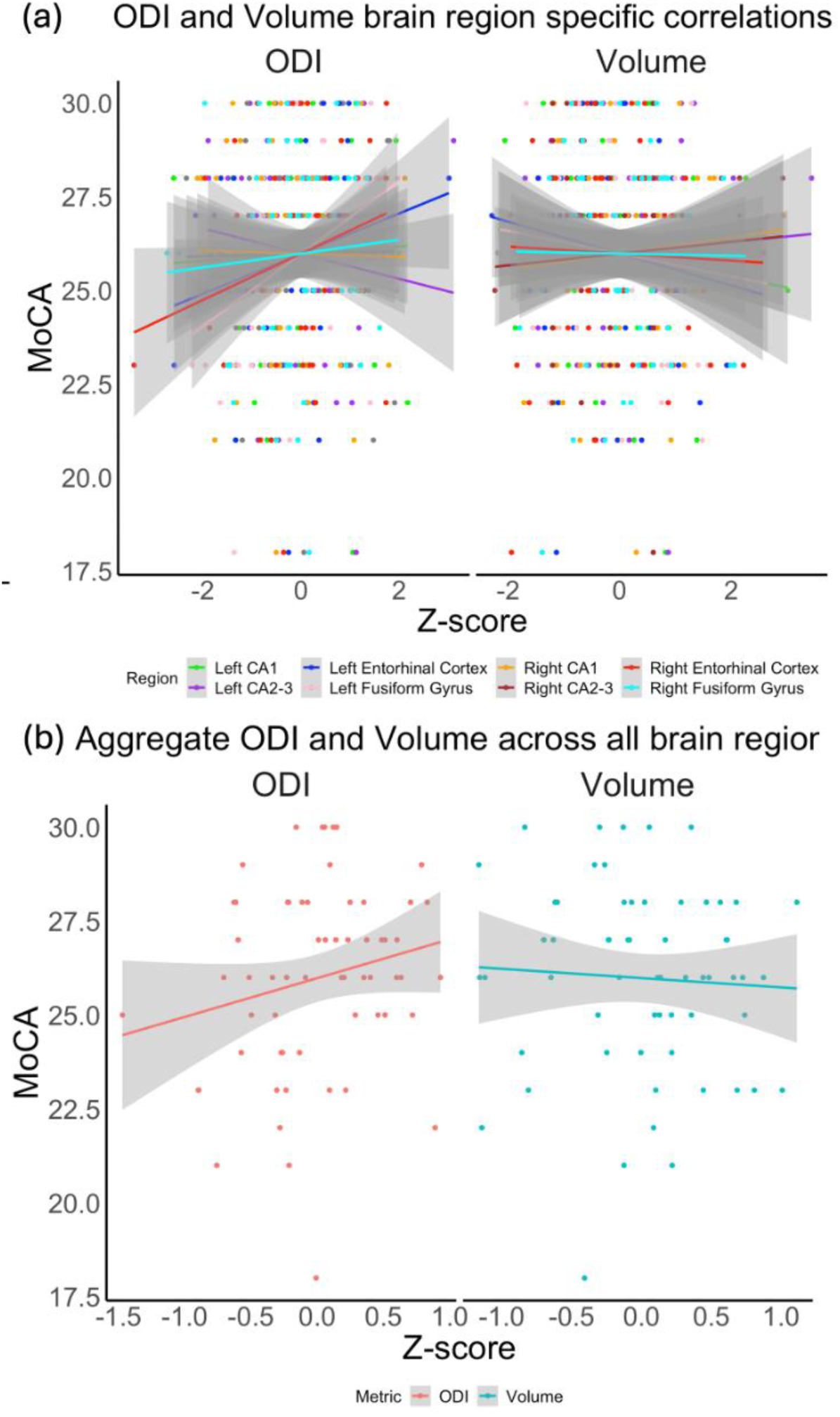
(a) Plots showing correlation between ODI and MoCA scores the left and right CA1, CA2-3, entorhinal cortex and fusiform gyrus. (b) Aggregate ODI and volume measures for each participant across all brain regions regressed to MoCA score.

**Table 2.**
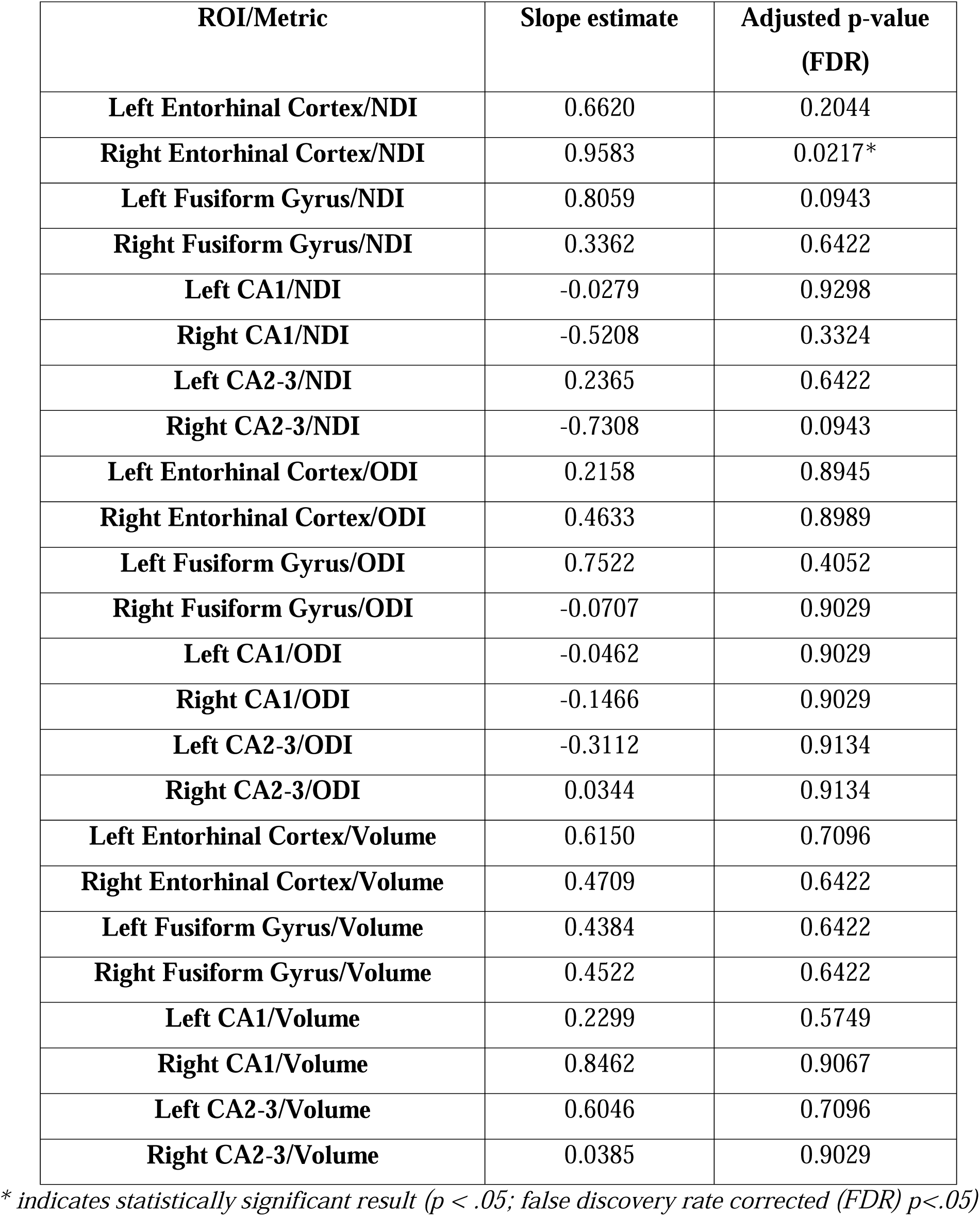
Individual relationship for each brain region based on either volume or NODDI metric quantification to MOCA. Raw and adjusted MOCA scores are reported here along with the estimated slope between each measure and MOCA. Adjusted p-value was calculated using a false discovery rate correction (FDR).

| **ROI/Metric** | **Slope estimate** | **Adjusted p-value**<br>**(FDR)** |
| --- | --- | --- |
| **Left Entorhinal Cortex/NDI** | 0.6620 | 0.2044 |
| **Right Entorhinal Cortex/NDI** | 0.9583 | 0.0217* |
| **Left Fusiform Gyrus/NDI** | 0.8059 | 0.0943 |
| **Right Fusiform Gyrus/NDI** | 0.3362 | 0.6422 |
| **Left CA1/NDI** | -0.0279 | 0.9298 |
| **Right CA1/NDI** | -0.5208 | 0.3324 |
| **Left CA2-3/NDI** | 0.2365 | 0.6422 |
| **Right CA2-3/NDI** | -0.7308 | 0.0943 |
| **Left Entorhinal Cortex/ODI** | 0.2158 | 0.8945 |
| **Right Entorhinal Cortex/ODI** | 0.4633 | 0.8989 |
| **Left Fusiform Gyrus/ODI** | 0.7522 | 0.4052 |
| **Right Fusiform Gyrus/ODI** | -0.0707 | 0.9029 |
| **Left CA1/ODI** | -0.0462 | 0.9029 |
| **Right CA1/ODI** | -0.1466 | 0.9029 |
| **Left CA2-3/ODI** | -0.3112 | 0.9134 |
| **Right CA2-3/ODI** | 0.0344 | 0.9134 |
| **Left Entorhinal Cortex/Volume** | 0.6150 | 0.7096 |
| **Right Entorhinal Cortex/Volume** | 0.4709 | 0.6422 |
| **Left Fusiform Gyrus/Volume** | 0.4384 | 0.6422 |
| **Right Fusiform Gyrus/Volume** | 0.4522 | 0.6422 |
| **Left CA1/Volume** | 0.2299 | 0.5749 |
| **Right CA1/Volume** | 0.8462 | 0.9067 |
| **Left CA2-3/Volume** | 0.6046 | 0.7096 |
| **Right CA2-3/Volume** | 0.0385 | 0.9029 |

\* indicates statistically significant result ( $p < .05$ ; false discovery rate corrected (FDR) $p < .05$ )

## 4. Discussion

This study used advanced diffusion MRI metric, in this case those derived using the NODDI model, to compare microstructural integrity between cognitively unimpaired APOE-e4 carriers and non-carriers in key brain regions associated with AD pathogenesis. As expected, neurite density (as measured by the NODDI-derived NDI parameter) was lower in the fusiform gyrus, entorhinal cortex and left hippocampal regions of carriers of the APOE-e4 allele when compared against non-carriers, although no such differences were observed in the right hippocampus. Results also showed that NDI values were sensitive to global cognition (measured as MoCA score) and APOE-e4 carrier status. Standard structural MRI-based volumetric measures showed no sensitivity to carrier status and ODI associated with carrier status only in the left and right CA1 regions. Neither volumetric measures nor ODI associated with MoCA performance. These results collectively support the hypothesis that, in individuals at genetic risk for developing AD, NDI is sensitive to early microstructural changes in brain regions that are selectively affected by AD, even before symptom onset.

Our results are reflective of and supported by several studies using the ADNI dataset, which have shown an association between early onset AD and NDI/ODI measures in the entorhinal, middle temporal, inferior temporal, fusiform, precuneus, and precentral areas^58–61^. Additionally, research outside of the ADNI dataset has validated NDI and ODI values in cortical regions like the fusiform gyrus and entorhinal cortex among participants with mild cognitive impairment (MCI) and AD, as well as similar age-related microstructural deficits ^26,58–61^. Our study significantly expands on these efforts by focusing on NODDI-measured microstructural deficits in people that are *cognitively normal* yet at genetic risk of developing AD (here, carriers of the APOE-e4 allele). Taken collectively, there is strong support of NDI as an early indicator of neurodegeneration. The fact that NDI can detect microstructural changes within brain regions that may eventually show marked pathology supports the use of advanced dMRI and biophysical modelling methods to longitudinally monitoring disease progression, particularly in the prodromal stages of AD.

While ODI was assessed in our study of cognitively normal subjects at genetic risk for AD, it was sensitive to APOE-e4 carrier status in fewer brain regions, and it did not associate with MoCA performance in our study. Since ODI measures the variability in neurite orientations (i.e., how dispersed or aligned the neurites are) it may, in theory, be more relevant as the disease progresses, when extensive disorganization and loss of neurite coherence become more evident due to more pronounced structural changes ^62–64^ and, ultimately, measurable differences using standard diffusion tensor imaging and tractography^14,65^.

Results from this study are consistent with prior work showing less connectivity of the left hippocampus with the rest of brain relative to the right hippocampus in cognitively unimpaired older adults who are APOE-e4 carriers^22,56^, and that the extent of left (but not right) hippocampal connectivity was correlated with verbal memory scores ^56^. Other studies ^21,35,57^ further support that AD progression affects the hippocampus asymmetrically, highlighting the critical role of left hemisphere microstructural differences linked to the APOE-e4 allele in early cognitive decline in at-risk individuals. This likely explains, in part, the differences in NDI values between APOE e4 carriers and non-carriers within the left but not right hippocampal subfields. This study does not, however, address whether microstructural changes within the left hippocampus precede or are concurrent with connectivity and cognitive changes, which can be explored in the future with longitudinal cohorts and rodent imaging studies.

A key limitation of this study lies in the degree to which the NODDI biophysical model accurately reflects non-white matter tissue microstructure. NODDI was originally “tuned” for assessment of white matter structures ^58,61,66^ and thus makes key model assumptions about the tissue that do not completely reflect the “mixed” nature of subcortical structures like the hippocampus (i.e., cell bodies, unmyelinated axons, and myelinated axons all existing within a single voxel), nor does it account for potential exchange of water across cellular compartments. Nevertheless, given the acquisition scheme of the dataset used, our focus on neurite microstructure, and the uncertain importance of exchange in biophysical models of diffusion in the brain given real-world clinical datasets ^67–69^, NODDI was deemed the most robust biophysical model for this preliminary study of subcortical microstructure in asymptomatic APOE e4 carrier and non-carrier groups. Still, the inability of NODDI to quantify microstructural detail about the soma (gray matter) or cellular exchange across tissue microenvironments, both of which may be critical to AD progression and treatment ^70–74^, is an important limitation of this work. Future work will focus on optimization of data acquisition parameters for more complex diffusion models such as soma and neurite density imaging (SANDI) ^75^ and/or neurite exchange imaging (NEXI) ^67^, which will capture and/or mitigate these potentially important parameters. Finally, the relatively small sample size, particularly of carriers, and the cross-sectional nature of this study limits the generalizability of the findings.

Future studies with larger cohorts are needed to confirm these results and enhance the statistical power of the analyses. Despite this limitation, the observed trends provide valuable insights into the potential impact of APOE-e4 status on very early microstructural changes to the brain that might be pathologically relevant to the progression of AD.

This study used NODDI analyses of diffusion MRI to show microstructural differences between cognitively intact APOE-e4 carriers and non-carriers, specifically in the fusiform gyrus, entorhinal cortex and left hippocampal subfields (CA1, CA2-3). These results are particularly promising, given that more conventional MRI methods (i.e., T1-weighted imaging) and cognitive screening tools (i.e., MoCA) were not sensitive to differences between these groups who have differential genetic risk for AD. This study therefore suggests that dMRI could be used to: (i) study AD progression at very early, asymptomatic stages, (ii) potentially stratify AD risk within asymptomatic individuals at early stages, and (iii) monitor disease progression over time, particularly in the prodromal stages of AD.

## Data Availability

All data produced are available online at adni.loni.usc.edu

https://adni.loni.usc.edu/

## Acknowledgments

Data collection and sharing for this project was funded by the Alzheimer’s Disease Neuroimaging Initiative (ADNI) (National Institutes of Health Grant U01 AG024904) and DOD ADNI (Department of Defense award number W81XWH-12-2-0012). ADNI is funded by the National Institute on Aging, the National Institute of Biomedical Imaging and Bioengineering, and through generous contributions from the following: AbbVie, Alzheimer’s Association; Alzheimer’s Drug Discovery Foundation; Araclon Biotech; BioClinica, Inc.; Biogen; Bristol- Myers Squibb Company; CereSpir, Inc.; Cogstate; Eisai Inc.; Elan Pharmaceuticals, Inc.; Eli Lilly and Company; EuroImmun; F. Hoffmann-La Roche Ltd and its affiliated company Genentech, Inc.; Fujirebio; GE Healthcare; IXICO Ltd.;Janssen Alzheimer Immunotherapy Research & Development, LLC.; Johnson & Johnson Pharmaceutical Research & Development LLC.; Lumosity; Lundbeck; Merck & Co., Inc.;Meso Scale Diagnostics, LLC.; NeuroRx Research; Neurotrack Technologies; Novartis Pharmaceuticals Corporation; Pfizer Inc.; Piramal Imaging; Servier; Takeda Pharmaceutical Company; and Transition Therapeutics. The Canadian Institutes of Health Research is providing funds to support ADNI clinical sites in Canada. Private sector contributions are facilitated by the Foundation for the National Institutes of Health (www.fnih.org). The grantee organization is the Northern California Institute for Research and Education, and the study is coordinated by the Alzheimer’s Therapeutic Research Institute at the University of Southern California. ADNI data are disseminated by the Laboratory for Neuro Imaging at the University of Southern California.

Dr. Scott Beeman and Dr. Sydney Schaefer are affiliated with the Arizona Alzheimer’s Consortium, and Dr. Schaefer is a REC Fellow of the Arizona Alzheimer’s Disease Research Center (P30 AG072980).

## Consent Statement

All human subjects participating in studies conducted by the Alzheimer’s Disease Neuroimaging Initiative (ADNI), provided informed consent.

## References

1. Zhang Y, Chen H, Li R, Sterling K, Song W. Amyloid β-based therapy for Alzheimer’s disease: challenges, successes and future. Signal Transduct Target Ther. 2023;8(1):1–26. doi:10.1038/s41392-023-01484-7

2. Albert SM, Glied S, Andrews H, Stern Y, Mayeux R. Primary care expenditures before the onset of Alzheimer’s disease. Neurology. 2002;59(4):573–578. doi:10.1212/WNL.59.4.573

3. Landau SM, Harvey D, Madison CM, et al. Associations between cognitive, functional, and FDG-PET measures of decline in AD and MCI. Neurobiol Aging. 2011;32(7):1207–1218. doi:10.1016/j.neurobiolaging.2009.07.002

4. Reiman EM, Caselli RJ, Chen K, Alexander GE, Bandy D, Frost J. Declining brain activity in cognitively normal apolipoprotein E 4 heterozygotes: A foundation for using positron emission tomography to efficiently test treatments to prevent Alzheimer’s disease. Proc Natl Acad Sci. 2001;98(6):3334–3339. doi:10.1073/pnas.061509598

5. Nordberg A, Rinne J, Kadir A, Langstrom B. The use of PET in Alzheimer disease. Nat Rev Neurol. 2010;6:78–87. doi:10.1038/nrneurol.2009.217

6. Saint-Aubert L, Lemoine L, Chiotis K, Leuzy A, Rodriguez-Vieitez E, Nordberg A. Tau PET imaging: present and future directions. Mol Neurodegener. 2017;12(1):19. doi:10.1186/s13024-017-0162-3

7. Mak E, Dounavi ME, Operto G, et al. APOE 4 exacerbates age-dependent deficits in cortical microstructure. Brain Commun. 2024;6(1):fcad351. doi:10.1093/braincomms/fcad351

8. Mukherji D, Mukherji M, Mukherji N, Alzheimer’s Disease Neuroimaging Initiative. Early detection of Alzheimer’s disease using neuropsychological tests: a predict–diagnose approach using neural networks. Brain Inform. 2022;9(1):23. doi:10.1186/s40708-022-00169-1

9. Albert MS, Moss MB, Tanzi R, Jones K. Preclinical prediction of AD using neuropsychological tests. J Int Neuropsychol Soc. 2001;7(5):631–639. doi:10.1017/S1355617701755105

10. Frisoni GB, Fox NC, Jack CR, Scheltens P, Thompson PM. The clinical use of structural MRI in Alzheimer disease. Nat Rev Neurol. 2010;6(2):67–77. doi:10.1038/nrneurol.2009.215

11. Goveas J, O’Dwyer L, Mascalchi M, et al. Diffusion-MRI in neurodegenerative disorders. Magn Reson Imaging. 2015;33(7):853–876. doi:10.1016/j.mri.2015.04.006

12. Douaud G, Menke RAL, Gass A, et al. Brain Microstructure Reveals Early Abnormalities more than Two Years prior to Clinical Progression from Mild Cognitive Impairment to Alzheimer’s Disease. J Neurosci. 2013;33(5):2147–2155. doi:10.1523/JNEUROSCI.4437-12.2013

13. Hoy AR, Ly M, Carlsson CM, et al. Microstructural white matter alterations in preclinical Alzheimer’s disease detected using free water elimination diffusion tensor imaging. PLOS ONE. 2017;12(3):e0173982. doi:10.1371/journal.pone.0173982

14. Zhuang L, Sachdev PS, Trollor JN, et al. Microstructural White Matter Changes, Not Hippocampal Atrophy, Detect Early Amnestic Mild Cognitive Impairment. PLOS ONE. 2013;8(3):e58887. doi:10.1371/journal.pone.0058887

15. Kamiya K, Hori M, Aoki S. NODDI in clinical research. J Neurosci Methods. 2020;346. doi:10.1016/j.jneumeth.2020.108908

16. Wen Q, Kelley DAC, Banerjee S, et al. Clinically feasible NODDI characterization of glioma using multiband EPI at 7 T. NeuroImage Clin. 2015;9:291–299. doi:10.1016/j.nicl.2015.08.017

17. Tournier JD. Diffusion MRI in the brain – Theory and concepts. Prog Nucl Magn Reson Spectrosc. 2019;112–113:1-16. doi:10.1016/j.pnmrs.2019.03.001

18. Yu X, Przybelski SA, Reid RI, et al. NODDI in gray matter is a sensitive marker of aging and early AD changes. Alzheimers Dement Diagn Assess Dis Monit. 2024;16(3):e12627. doi:10.1002/dad2.12627

19. Xu Y, Jack CR, O’Brien PC, et al. Usefulness of MRI measures of entorhinal cortex versus hippocampus in AD. Neurology. 2000;54(9):1760–1767. doi:10.1212/WNL.54.9.1760

20. Zhang H, Schneider T, Wheeler-Kingshott CA, Alexander DC. NODDI: Practical in vivo neurite orientation dispersion and density imaging of the human brain. NeuroImage. 2012;61(4):1000–1016. 10.1016/j.neuroimage.2012.03.072

21. Abraham M, Seidenberg M, Kelly DA, et al. Episodic Memory and Hippocampal Volume Predict Five-Year MCI Conversion in Healthy APOE ε4 Carriers. J Int Neuropsychol Soc JINS. 2020;26(7):733–738. doi:10.1017/S1355617720000181

22. Caselli RJ, Reiman EM, Osborne D, et al. Longitudinal changes in cognition and behavior in asymptomatic carriers of the APOE e4 allele. Neurology. 2004;62(11):1990–1995. doi:10.1212/01.WNL.0000129533.26544.BF

23. van Duijn CM, de Knijff P, Cruts M, et al. Apolipoprotein E4 allele in a population–based study of early–onset Alzheimer’s disease. Nat Genet. 1994;7(1):74–78. doi:10.1038/ng0594-74

24. Locke PA, Conneally PM, Tanzi RE, Gusella JF, Haines JL. Apolipoprotein E4 allele and Alzheimer disease: Examination of Allelic association and effect on age at onset in both early and late onset cases. Genet Epidemiol. 1995;12(1):83–92. doi:10.1002/gepi.1370120108

25. Evans SL, Dowell NG, Prowse F, Tabet N, King SL, Rusted JM. Mid age APOE ε4 carriers show memory-related functional differences and disrupted structure-function relationships in hippocampal regions. Sci Rep. 2020;10(1):3110. doi:10.1038/s41598-020-59272-0

26. Du J, Liu Z, Hanford LC, et al. Exploration of Alzheimer’s Disease MRI Biomarkers Using APOE4 Carrier Status in the UK Biobank. Published online September 14, 2021:2021.09.09.21263324. doi:10.1101/2021.09.09.21263324

27. Cortical microstructural alterations along the Alzheimer’s disease continuum and association with amyloid and tau pathology. doi:10.21203/rs.3.rs-3921380/v1

28. Slattery CF, Zhang J, Paterson RW, et al. ApoE influences regional white-matter axonal density loss in Alzheimer’s disease. Neurobiol Aging. 2017;57:8–17. doi:10.1016/j.neurobiolaging.2017.04.021

29. Burggren A, Brown J. Imaging markers of structural and functional brain changes that precede cognitive symptoms in risk for Alzheimer’s disease. Brain Imaging Behav. 2014;8(2):251–261. doi:10.1007/s11682-013-9278-4

30. Mak E, Reid RI, Przybelski SA, et al. Alzheimer’s disease biomarkers of amyloid and tau influence white matter neurite alterations in dementia with Lewy bodies: a NODDI study. Alzheimers Dement. 2023;19(S16):e076922. doi:10.1002/alz.076922

31. Twamley EW, Ropacki S a. L, Bondi MW. Neuropsychological and neuroimaging changes in preclinical Alzheimer’s disease. J Int Neuropsychol Soc. 2006;12(5):707–735. doi:10.1017/S1355617706060863

32. Devanand DP, Bansal R, Liu J, Hao X, Pradhaban G, Peterson BS. MRI hippocampal and entorhinal cortex mapping in predicting conversion to Alzheimer’s disease. NeuroImage. 2012;60(3):1622–1629. doi:10.1016/j.neuroimage.2012.01.075

33. Malek-Ahmadi M, Duff K, Chen K, et al. Volumetric regional MRI and neuropsychological predictors of motor task variability in cognitively unimpaired, Mild Cognitive Impairment, and probable Alzheimer’s disease older adults. Exp Gerontol. 2023;173:112087. doi:10.1016/J.EXGER.2023.112087

34. Hamza, Eid A., Moustafa, Ahmed A., Tindle, Richard, et al. Effect of APOE4 Allele and Gender on the Rate of Atrophy in the Hippo-campus, Entorhinal Cortex, and Fusiform Gyrus in Alzheimer’s Disease. Curr Alzheimer Res. 2022;19:943–953. doi:10.2174/1567205020666230309113749

35. Jessen F, Feyen L, Freymann K, et al. Volume reduction of the entorhinal cortex in subjective memory impairment. Neurobiol Aging. 2006;27(12):1751–1756. doi:10.1016/j.neurobiolaging.2005.10.010

36. Ma D, Fetahu IS, Wang M, et al. The fusiform gyrus exhibits an epigenetic signature for Alzheimer’s disease. Clin Epigenetics. 2020;12(1):129. doi:10.1186/s13148-020-00916-3

37. Weiner MW, Veitch DP, Aisen PS, et al. The Alzheimer’s Disease Neuroimaging Initiative 3: continued innovation for clinical trial improvement. Alzheimers Dement J Alzheimers Assoc. 2017;13(5):561–571. doi:10.1016/j.jalz.2016.10.006

38. Jenkinson M, Beckmann CF, Behrens TEJ, Woolrich MW, Smith SM. FSL. NeuroImage. 2012;62(2):782-790. doi:10.1016/j.neuroimage.2011.09.015

39. Tournier JD, Smith R, Raffelt D, et al. MRtrix3: A fast, flexible and open software framework for medical image processing and visualisation. NeuroImage. 2019;202:116137. doi:10.1016/j.neuroimage.2019.116137

40. Schilling KG, Blaber J, Huo Y, et al. Synthesized b0 for diffusion distortion correction (Synb0-DisCo). Magn Reson Imaging. 2019;64:62–70. doi:10.1016/J.MRI.2019.05.008

41. Andersson JLR, Sotiropoulos SN. An integrated approach to correction for off-resonance effects and subject movement in diffusion MR imaging. NeuroImage. 2016;125:1063–1078. doi:10.1016/j.neuroimage.2015.10.019

42. Microstructure Imaging Group | Download NODDI matlab toolbox. Accessed January 25, 2024. http://mig.cs.ucl.ac.uk/index.php?n=Download.NODDI

43. Jenkinson M, Bannister P, Brady M, Smith S. Improved optimization for the robust and accurate linear registration and motion correction of brain images. NeuroImage. 2002;17(2):825–841. doi:10.1016/s1053-8119(02)91132-8

44. Andersson JLR. Non-linear registration aka Spatial normalisation. Accessed March 7, 2024. https://www.fmrib.ox.ac.uk/datasets/techrep/tr07ja2/tr07ja2.pdf

45. https://www.bic.mni.mcgill.ca/ServicesAtlases/HomePage. Accessed March 7, 2024. https://www.bic.mni.mcgill.ca/ServicesAtlases/HomePage

46. Desikan RS, Ségonne F, Fischl B, et al. An automated labeling system for subdividing the human cerebral cortex on MRI scans into gyral based regions of interest. NeuroImage. 2006;31(3):968–980. doi:10.1016/j.neuroimage.2006.01.021

47. Gordon BA, Zacks JM, Blazey T, et al. Task-evoked fMRI changes in attention networks are associated with preclinical Alzheimer’s disease biomarkers. Neurobiol Aging. 2015;36(5):1771–1779. doi:10.1016/j.neurobiolaging.2015.01.019

48. Yetkin FZ, Rosenberg RN, Weiner MF, Purdy PD, Cullum CM. FMRI of working memory in patients with mild cognitive impairment and probable Alzheimer’s disease. Eur Radiol. 2006;16(1):193–206. doi:10.1007/s00330-005-2794-x

49. Berghuis KMM, Fagioli S, Maurits NM, et al. Age-related changes in brain deactivation but not in activation after motor learning. NeuroImage. 2019;186:358–368. doi:10.1016/j.neuroimage.2018.11.010

50. Bo J, Seidler RD. Visuospatial Working Memory Capacity Predicts the Organization of Acquired Explicit Motor Sequences. J Neurophysiol. 2009;101(6):3116–3125. doi:10.1152/jn.00006.2009

51. Langan J, Seidler RD. Age differences in spatial working memory contributions to visuomotor adaptation and transfer. Behav Brain Res. 2011;225(1):160–168. doi:10.1016/j.bbr.2011.07.014

52. R: The R Project for Statistical Computing. Accessed January 25, 2024. https://www.r-project.org/

53. Killiany RJ, Hyman BT, Gomez-Isla T, et al. MRI measures of entorhinal cortex vs hippocampus in preclinical AD. Neurology. 2002;58(8):1188–1196. doi:10.1212/WNL.58.8.1188

54. Rao YL, Ganaraja B, Murlimanju BV, Joy T, Krishnamurthy A, Agrawal A. Hippocampus and its involvement in Alzheimer’s disease: a review. 3 Biotech. 2022;12(2):55. doi:10.1007/s13205-022-03123-4

55. Pennanen C, Kivipelto M, Tuomainen S, et al. Hippocampus and entorhinal cortex in mild cognitive impairment and early AD. Neurobiol Aging. 2004;25(3):303–310. doi:10.1016/S0197-4580(03)00084-8

56. Baxter LC, Limback-Stokin M, Patten KJ, et al. Hippocampal connectivity and memory decline in cognitively intact APOE ε4 carriers. Alzheimers Dement. 2023;19(9):3806–3814. doi:10.1002/alz.13023

57. Mohtasib R, Alghamdi J, Jobeir A, et al. MRI biomarkers for Alzheimer’s disease: the impact of functional connectivity in the default mode network and structural connectivity between lobes on diagnostic accuracy. Heliyon. 2022;8(2):e08901. doi:10.1016/j.heliyon.2022.e08901

58. Parker TD, Slattery CF, Zhang J, et al. Cortical microstructure in young onset Alzheimer’s disease using neurite orientation dispersion and density imaging. Hum Brain Mapp. 2018;39(7):3005–3017. doi:10.1002/hbm.24056

59. Colgan N, Siow B, O’Callaghan JM, et al. Application of neurite orientation dispersion and density imaging (NODDI) to a tau pathology model of Alzheimer’s disease. NeuroImage. 2016;125:739–744. doi:10.1016/j.neuroimage.2015.10.043

60. Fu X, Shrestha S, Sun M, et al. Microstructural White Matter Alterations in Mild Cognitive Impairment and Alzheimer’s Disease. Clin Neuroradiol. 2020;30(3):569–579. doi:10.1007/s00062-019-00805-0

61. Billiet T, Vandenbulcke M, Mädler B, et al. Age-related microstructural differences quantified using myelin water imaging and advanced diffusion MRI. Neurobiol Aging. 2015;36(6):2107–2121. doi:10.1016/j.neurobiolaging.2015.02.029

62. Jack CR, Holtzman DM. Biomarker Modeling of Alzheimer’s Disease. Neuron. 2013;80(6):1347–1358. doi:10.1016/j.neuron.2013.12.003

63. Weston PSJ, Coath W, Harris MJ, et al. Cortical tau is associated with microstructural imaging biomarkers of neurite density and dendritic complexity in Alzheimer’s disease. Alzheimers Dement. 2023;19(6):2750–2754. doi:10.1002/alz.13011

64. Vogt NM, Hunt JF, Adluru N, et al. Cortical Microstructural Alterations in Mild Cognitive Impairment and Alzheimer’s Disease Dementia. Cereb Cortex. 2020;30(5):2948–2960. doi:10.1093/cercor/bhz286

65. Jeurissen B, Leemans A, Tournier JD, Jones DK, Sijbers J. Investigating the prevalence of complex fiber configurations in white matter tissue with diffusion magnetic resonance imaging. Hum Brain Mapp. 2013;34(11):2747–2766. doi:10.1002/hbm.22099

66. Pines AR, Cieslak M, Larsen B, et al. Leveraging multi-shell diffusion for studies of brain development in youth and young adulthood. Dev Cogn Neurosci. 2020;43:100788. doi:10.1016/j.dcn.2020.100788

67. Jelescu IO, de Skowronski A, Geffroy F, Palombo M, Novikov DS. Neurite Exchange Imaging (NEXI): A minimal model of diffusion in gray matter with inter-compartment water exchange. NeuroImage. 2022;256:119277. doi:10.1016/J.NEUROIMAGE.2022.119277

68. Yang DM, Huettner JE, Bretthorst GL, Neil JJ, Garbow JR, Ackerman JJH. Intracellular Water Preexchange Lifetime in Neurons and Astrocytes. Magn Reson Med. 2018;79(3):1616–1627. doi:10.1002/mrm.26781

69. Schiavi S, Palombo M, Zacà D, et al. Mapping tissue microstructure across the human brain on a clinical scanner with soma and neurite density image metrics. Hum Brain Mapp. 2023;44(13):4792–4811. doi:10.1002/hbm.26416

70. Jelescu IO, Palombo M, Bagnato F, Schilling KG. Challenges for biophysical modeling of microstructure. J Neurosci Methods. 2020;344:108861. doi:10.1016/j.jneumeth.2020.108861

71. Nazeri A, Schifani C, Anderson JAE, Ameis SH, Voineskos AN. In Vivo Imaging of Gray Matter Microstructure in Major Psychiatric Disorders: Opportunities for Clinical Translation. Biol Psychiatry Cogn Neurosci Neuroimaging. 2020;5(9):855–864. doi:10.1016/j.bpsc.2020.03.003

72. Gatto RG. Molecular and microstructural biomarkers of neuroplasticity in neurodegenerative disorders through preclinical and diffusion magnetic resonance imaging studies. J Integr Neurosci. 2020;19(3):571–592. doi:10.31083/j.jin.2020.03.165

73. Figini M. Model-based analysis of diffusion Magnetic Resonance: study of microstructural damage in white matter and gray matter diseases. Published online September 30, 2014. Accessed August 5, 2024. https://www.politesi.polimi.it/handle/10589/97955

74. Singh K, Barsoum S, Schilling KG, An Y, Ferrucci L, Benjamini D. Neuronal microstructural changes in the human brain are associated with neurocognitive aging. Aging Cell. 2024;23(7):e14166. doi:10.1111/acel.14166

75. Palombo M, Ianus A, Guerreri M, et al. SANDI: A compartment-based model for non-invasive apparent soma and neurite imaging by diffusion MRI. NeuroImage. 2020;215:116835. doi:10.1016/J.NEUROIMAGE.2020.116835

